# Comparison of Bayesian methods for incorporating adult clinical trial data to improve certainty of treatment effect estimates in children

**DOI:** 10.1101/2023.02.02.23285367

**Authors:** R Walker, B Phillips, S Dias

## Abstract

There are challenges associated with recruiting children to take part in randomised clinical trials and as a result, compared to adults, in many disease areas we are less certain about which treatments are most safe and effective. This can lead to weaker recommendations about which treatments to prescribe in practice. However, it may be possible to ‘borrow strength’ from adult evidence to improve our understanding of which treatments work best in children, and many different statistical methods are available to conduct these analyses. In this paper we discuss Bayesian methods for extrapolating adult clinical trial evidence to children. Using an exemplar dataset, we compare the effect of modelling assumptions on the estimated treatment effect and associated heterogeneity. We finally discuss the appropriateness of different modelling assumptions in the context of estimating treatment effect in children.

## Introduction

There are various challenges associated with conducting randomised control trials (RCTs) to compare the efficacy and safety of medical interventions in children. A lower disease incidence in children means fewer patients are eligible to take part in clinical trials and research groups and pharmaceutical companies are wary of the increased effort which is required to conduct research with this population. Further to this, young people and/or their parents/carers may not wish to take on the additional burden and time commitment associated with being in a clinical trial (1) (2).

Despite many diseases affecting both adults and children, these challenges mean that when compared to adults, there are usually fewer RCTs in children and therefore, a greater uncertainty about which medicines work ‘best’ to treat a particular disease or condition. This can lead to medicines being licenced (authorised for marketing on the basis of quality, safety and efficacy) in adults some years before they become available for children (2). Prescribers may also have little alternative but to use medicines off-label without having direct information to inform their decision. This in turn, can create inconsistency in which medicines are prescribed to children to treat a particular condition and may mean that not all children are receiving the most effective treatment available.

To inform decision making within healthcare (including which medicines to prescribe for a condition), RCT evidence is often combined using evidence synthesis techniques, such as meta-analysis (MA), that take a weighted average of efficacy or safety results from multiple clinical trials (or studies) to produce a summary estimate of the comparative or relative efficacy (the effect of one treatment compared to another) of two interventions (3). An extension of this technique is a network-meta-analysis (NMA), sometimes referred to as a mixed-treatment comparison, that allows for synthesis of three or more treatments and the simultaneous comparison of each treatment with every other. NMA can also provide ‘indirect comparisons’ through which estimates of the relative efficacy or safety of treatments that have not been compared in head-to-head clinical trials are produced, provided they are present in a ‘connected network’ of treatments i.e., a network where there is a path between any two interventions with paths formed of randomised comparisons (4).

In paediatric research, however, these analyses may still not provide sufficient evidence for healthcare decision making as the required RCT evidence may be from a small number of patients and form only a sparse or disconnected network of comparisons, meaning the pooled treatment effect remains uncertain. To overcome these issues, it may be possible to ‘extrapolate’ or ‘borrow strength’ from clinical trial evidence in a separate but related population. For the paediatric population, this could be an adult population. Information from the adult population would be extended to make inference about the efficacy of treatments in children and may reduce the uncertainty of treatment effect estimates in children. To justify this type of extrapolation, the disease manifestation and progression of the disease of interest, along with the exposure response relationship (that is the observed effects of a treatment at different doses) should already be established in children and the similarities and/or differences with the adult population understood (5).

If these conditions are satisfied, ‘borrowing of strength’ from the adult efficacy data can be facilitated by an extension of Bayesian MA or NMA, referred to as a Bayesian information sharing model (**ISM**). Working within a Bayesian framework allows you to combine prior information (e.g., data from adult clinical trials or clinician’s perspective) about a parameter (e.g., the relative treatment effect), with that from a new study, to produce a ‘posterior probability distribution’ i.e. the revised or updated probability of an event occurring after taking into consideration new information (6). If no specific prior information is available, then a vague prior distribution can be specified in the ISM.

In order to ‘borrow strength’ from the adult population, ISMs need to make certain assumptions about the relationship between adult and paediatric populations, in terms of the clinical efficacy of the treatment (7). This can range from clinical trial information of the adult population being completely generalisable to the paediatric population, where full information sharing would be appropriate, to the clinical trial information of the two populations being completely independent of one another and no information sharing can take place. The extent to which strength is borrowed from one population to another is then determined by the modelling assumptions, the precision of the data available for the different populations, and the extent of agreement across data sources (7).

In this paper, we use an exemplar dataset of 16 RCTs comparing two anti-sickness regimens for the prevention chemotherapy induced nausea and vomiting, to investigate how the modelling assumptions of four different ISMs impact the treatment effect estimates and associated heterogeneity in children. The ISMs compared in this paper, extend the traditional MA and we discuss the appropriateness of their assumptions in the context of estimating treatment effect in children.

### Exemplar data set

Information sharing methods were motivated by an exemplar dataset containing outcomes from 16 RCTs (four from paediatric populations and 12 from adult populations) comparing aprepitant (a newer antiemetic) or fosaprepitant (the intravenous version of aprepitant) with a control regimen of a 5HT3 antagonist + dexamethasone, for the treatment of chemotherapy induced nausea and vomiting.

To create this dataset, we identified RCTs from a recent clinical antiemetic guideline from the Paediatric Oncology Group of Ontario (POGO) (8) and the Multinational Association for Supportive Care in Cancer (MASCC) (9). Data were commonly reported as the proportion of patients experiencing a ‘complete response’ i.e., no vomiting, from the point of chemotherapy administration, up to five days afterwards. These data were extracted and converted into the number of participants *experiencing* a vomiting event in the treatment and in the control arm. A table of study characteristics and extracted outcome data is provided in the supporting information S1.

As the incidence of cancer in children is much rarer than in adults, but the therapeutic chemotherapies used are often the same, and the effects of antiemetic medications are clinically believed to be similar, it was considered appropriate to explore information sharing methods for this example.

## Methods

### The MA model

In the standard MA model for each study i and each arm k, the binary data in the form of events *r*_*ik*_ and total number of patients *n*_*ik*_, are described as coming from a binomial likelihood:

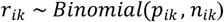

where *p*_*ik*_ is the probability of an event in arm k of study i, modelled on the log-scale and defining the linear predicator θ_ik_ i.e., the log risk of an event in arm k of trial i:

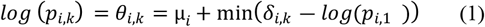

where μ_*i*_ is the study specific baseline treatment effects (i.e., the baseline risk of an outcome, which must be positive) in arm 1 of trial i, defined as μ_*i*_ = *log*(*p*_*i*, 1)_ where *p*_*i*,1_ is given a vague prior distribution (Uniform(0,1)), *δ*_*i,k*_ is the study specific relative treatment effect (i.e. the log risk ratio) of the treatment in arm k compared to the treatment in arm 1 *in that trial* where the relative effect of a treatment compared to itself, is set to zero: *δ*_*i*,1_ = 0.

Models can be specified as having a common/fixed effect, where it is assumed that all studies estimate the same relative effect i.e. the relative effect is not expected to differ between study populations included in the analysis; or models may have random effects, where relative effects of studies are assumed exchangeable or similar, i.e. the relative effect is expected to differ between study populations included in the analysis, but is expected to fall within a certain range or distribution (3).

For random effects model the study specific relative treatment effects are assumed to be drawn from a normal distribution with a common mean and between trial variability (i.e., the heterogeneity parameter) *τ*^2^. As we are only comparing two treatments in our application the common mean *d*_1,2_, represents the pooled relative effect of the treatment regimen 2 (aprepitant/fosaprepitant + a 5HT_3_ antagonist + dexamethasone) compared to the comparator treatment regimen 1 (a 5HT_3_ antagonist + dexamethasone) :

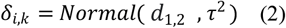

The parameter *d*_1,2_ is to be estimated and given a non-informative prior: *d*_1,2_ *∼* Normal(0,100^2^). For the fixed effects model equation 2 is replaced by:

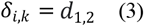

### Bayesian information sharing models

We now describe the four ISMs compared in this paper. These ISMs differ in parameters in which they share information: either relative treatment effects, or between study heterogeneity (for random effects models). As a result, different assumptions are made about the relationship between the two populations and different amounts of information are shared between them.

N.B ISMs that relate the parameters of the evidence sets using an exchangeability-based relationship (where a common distribution is imposed on the parameters e.g., in the multilevel models and the random walk model) are most useful when there are multiple sources of evidence (7). As we are only considering two populations, these models are not discussed in this paper.

### The splitting model

The first model we implement is the splitting model, which simply extends the standard MA model to accommodate the inclusion of two evidence sets but estimates model parameters separately for each population *j* (7).

For the random effect model, equation 2 is edited to include population specific parameters:

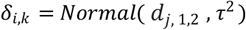

where *j* defines the population. *j =* 1 when study index *i* = 1,…, nsA (indicating the adult studies), and *j=* 2 when *i* = nsA + 1,…., nsA + nsC (indicating the children studies), where nsA is the number of adult studies and nsC is the number of children studies.

For the fixed effects model, equation 3 is edited to include population specific parameters:

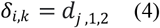

The treatment effects for population *j* are given vague priors: *d*_*j,1,2*_*∼* Normal(0,100^2^).

As such, one MA is performed on the adult data, and another on data from the children, although both analyses are conducted simultaneously. In the random effects model, information on between-study heterogeneity is shared between populations and is assumed the same for adults and children i.e., the variation in treatment effect between clinical trials is assumed the same in the adult and paediatric populations (although other assumptions could also be imposed e.g., that heterogeneity in children is proportional to adults). This produces a marginal benefit, in terms of supporting the estimation of treatment effect in children, as when there are few children studies there is little information about the variation in treatment effect between studies.

The fixed effect version of this model does not share any information between the populations, rather it estimates the treatment effects separately in adults and children, assuming they are completely independent of each other (see equation 4).

### Functional relationship models

This ISM assumes parameters are related using a deterministic function i.e. assuming that the relative effect in one population can be written as a function of the relative effect in the other (7):

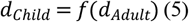

Where *d*_*Child*_ is the parameter that relates to the children’s evidence and *d*_*Adult*_ the parameter that relates to the adult evidence. The function f () can take different forms. Here we explore two relationships.

***The ‘lumping’ model***, ‘lumps’ the data together from the evidence sets and therefore assumes the adult data is completely generalisable to the children i.e., the function in equation 5 is the identity function and there is no difference in the relative effect between the two populations. This is equivalent to not distinguishing between adult and paediatric data and carrying out a simple meta-analysis using all the data.

***The proportional effects model***, assumes that the relative risk estimated in children is proportional to the relative risk estimated in the adult population, so that there is an additive relationship on the log relative risk scale:

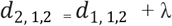

A vague prior distribution is specified for the relative treatment effect in adults on the log relative risk scale *d*_1, 1,2_ ∼ (Normal(0,100^2^)) and the change (in the log-RR scale in children) is measured by a parameter lambda (λ), which is also given a vague prior distribution λ∼ (Normal(0,100^2^)) (although an informative prior could be used for λ if appropriate). In this model, information sharing takes place from adult to children but also vice-versa. An alternative way of implementing this model is through modifying the BUGS code to include of a ‘cut’ function (4)(see supporting information S2).

### Meta-regression

An alternative model is where the two populations are indicated by a binary study-level covariate and a regression parameter i.e., the difference in treatment effect between the two populations, is estimated. Here the linear predicator (equation 1) is modified as follows (10):

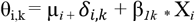

Where β_*1k*_ is the covariate effect of the indirect or (or adult) population X_i_ on the treatment effect. This model allows for a test of interaction between the relative treatment effect and the binary study level covariate that identifies the adult population. This will assess whether the relative treatment effect is dependent on data arising from the adult or children’s population. In this model, information sharing only occurs in random effects models where the heterogeneity is assumed equal across populations. In a fixed effect model this is equivalent to a subgroup analysis.

### Implementation

All analyses were carried out in OpenBUGS version 3.2.2 (11) and code to implement all models can be found in the supporting information S2. Whilst we include only one treatment comparison in our analyses (equivalent to a pairwise meta-analysis) and only two arm trials, the code provided in the supporting information S2 will accommodate multiple treatment comparisons and studies with three or more arms whilst adequately accounting for multiple random effects that are correlated (see (4)).

Separate MA models were conducted for adults and for children, followed by the four ISMs. For all models, vague prior distributions were used for all trial baselines and for relative treatment effects (Normal(0,100^2^)). For random effects models, a minimally informative prior distribution (Uniform(0,2)) was used for the between-study heterogeneity parameter. Results are based on 50,000 interactions on three chains after a burn-in of 10,000. Convergence was assessed visually by checking the mixing of chains. Model fit statistics including the deviance information criteria, and total residual deviance from each ISM model, along with treatment effect estimates and associated heterogeneity (and 95% credible intervals) were compared to those from the separately applied NMAs for adults and for children.

## Results

Table 1 presents treatment effect estimates and model fit statistics from both fixed and random effects models analysing data from adults and children’s populations separately, and together, using the four proposed models. The ‘lumping’ model, which assume relative effects in both populations are the same, produced treatment effect estimates closer to those of the MA using adult only data and had a larger between study heterogeneity compared to the other models, suggesting heterogeneity might be explained by assuming a less restrictive relationship for the relative effects across the populations.

**Table 1.**
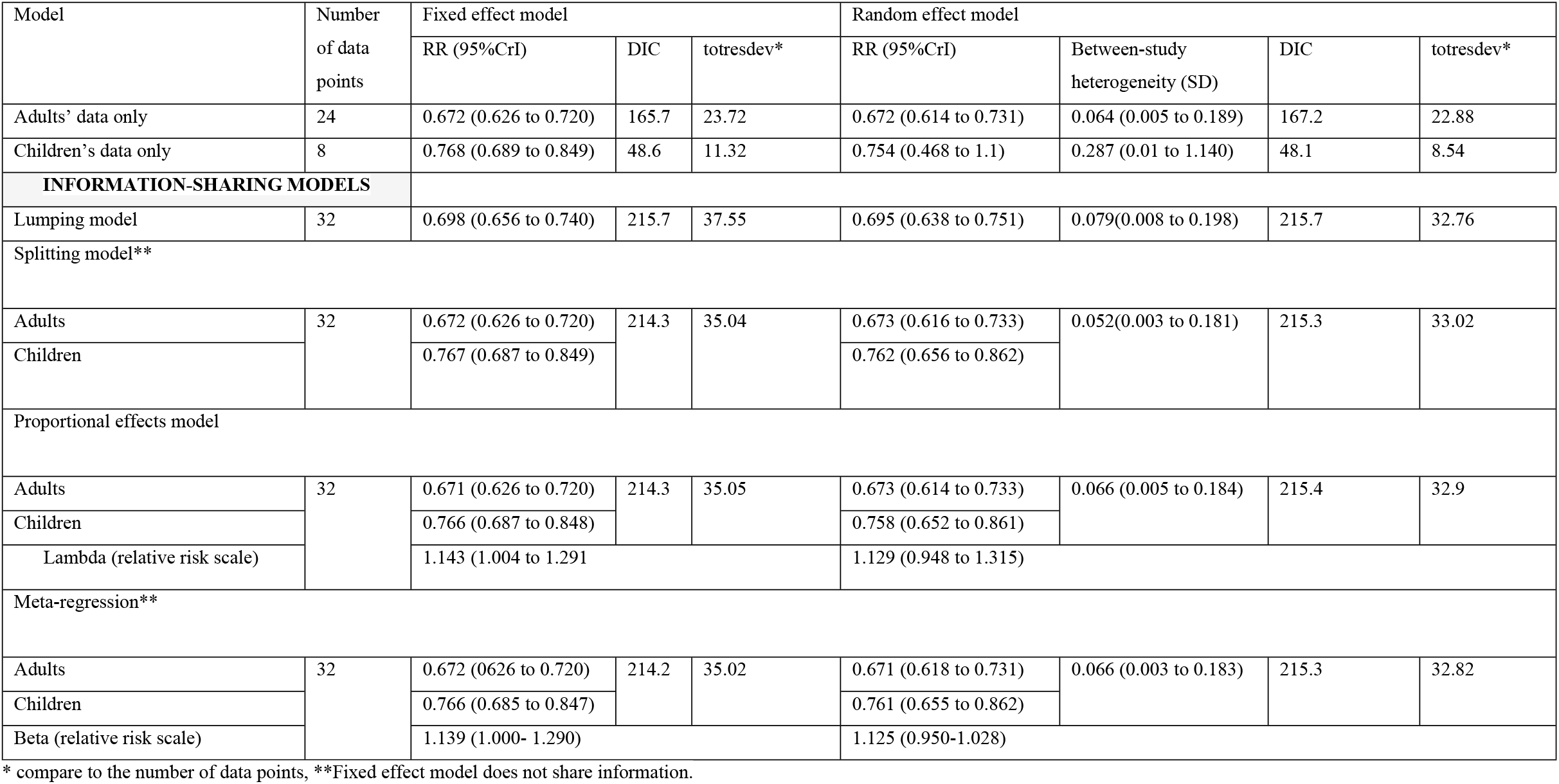
Results from MA and ISM models comparing aprepitant or fosaprepitant to control regimen for the treatment of chemotherapy induced nausea and vomiting.

The splitting model (with shared heterogeneity when random effects are considered), proportional effects and meta-regression model, produced estimates for adults and children comparable to the MAs conducted separately for the populations and the precision of the treatment effect estimate for children produced by the random effects model was improved in all models. Although there was little change in precision in fixed effect models, model fit was slightly improved (residual deviances were closer to the number of data points) compared to the lumping model. Across all information sharing models, fixed effects and random effects models produced comparable DICs. The total residual deviance was marginally lower for the random effects models (table 1), although differences were small meaning that in general the simplest of the two models (i.e., the fixed effect model with no additional parameters – ‘lumping model’) would be selected.

Lamba and beta estimates, indicating the difference between the treatment effect estimates in children and adults are comparable, with both models estimating a 14% and 13% increase in the relative risk for children compared to adults, for the fixed and random effects models respectively, although the 95% CrI for the latter includes the possibility of no difference in effects across the populations for both models (table 1).

## Discussion

In situations where the disease manifestation and progression, along with the exposure response relationship of a treatment is understood in children, ISMs could help to overcome the scarcity of clinical effectiveness evidence in this population, by including adult data into analyses(5). As ISMs may improve the precision or certainty of the relative treatment effect estimates in children, the analyses could maximise the usefulness of an existing evidence base in children to inform decisions about which treatment to prescribe for paediatric diseases and or illnesses. In this work we have compared the relative treatment effects and associated heterogeneity produced by different ISMs that incorporated adult data, to estimates from a MA using only data from children.

We found that when treatment effect estimates for children produced by the model that does not distinguish between populations (‘lumping model’) were compared to those produced by the MAs conducted separately for children, the estimates were closer to those from the adults MA. This is because there are more studies in the adult population and therefore the results are dominated by the adult information. In the random effect model the between study-heterogeneity was greater than that of the other information sharing models (Table 1). This is due to the assumption that the relative treatment effect and heterogeneity are equal for both populations and therefore the model needs to account for the additional heterogeneity resulting from pooling adult and paediatric data. Thus, we consider the ‘lumping model’ is likely not an appropriate choice, particularly when there is potential for differences in relative treatment effect between adults and children, as pooled estimates produced for children may not reflect the true effectiveness of the treatment in this population.

The ‘splitting’ model produced very similar treatment effect estimates to the MAs conducted using only children’s data. For the fixed effect model, this is expected as the model does not share information between the populations. While perhaps safest, in terms of not mistakenly assuming similarity in clinical efficacy, this maintains the status-quo of evidence sparsity for children. The random effect model shares information on the between study heterogeneity and therefore estimated this parameter in children more precisely than when the MA is conducted separately for children (Table 1), as it is able to borrow information from the adult data to better estimate the parameter. We note that the plausibility of this assumption (in our example, that the spread of effects in antiemetic trials in children would be the same as seen in adults), would need to be considered by paediatrics oncology experts. If considered appropriate, this model may be useful for reducing the uncertainty around treatment effect estimates when there are very few RCTs in children and a random effects model is considered appropriate.

The proportional effects model, which estimates the difference (λ) between the treatment effects across the two populations, produced comparable relative effects estimates to the MAs containing only children’s data (Table 1). This model is particularly advantageous, as it is able to include the adult data to support the sample size in the paediatric population, without assuming that the relative effectiveness is identical. This may be the ‘safer’ option when extrapolating from adult clinical trial data, as the efficacy of medications may vary from that of children due to differences in the way that medicines are absorbed, distributed, metabolised, and excreted in the body (2) (12).

However, in the proportional effects model, information sharing takes place from adult to children but also vice-versa, which may not be desirable. When the model was modified to prevent the data from the children affecting the estimate of the adult population, results were comparable to the MAs containing only data from children (see Supporting information S3). This modification may be helpful when adult data are particularly abundant and it is, therefore, not necessary, or desirable for the children’s data to influence adult relative effect estimates or decisions already made for adults.

Finally, the meta-regression model that estimates the effect of a binary covariate (adult/child) on the treatment outcome, again produced comparable results to the MAs containing only children’s data and produced very similar estimates to the proportional effects model. This is expected as both models are estimating conceptually similar parameters in different ways, the proportional effects model estimates λ as an unknown parameter from the treatment effect estimates, whilst the meta-regression model includes the adult/children as a binary study-level covariate in the linear predictor, then estimates the difference in treatment effect between adults and children. The meta-regression model may be particularly useful to explore further differences in treatment effect between adults and children. The model can be extended to include baseline risk or other covariate in the model, to understand whether differences are attributable to differences in underlying risk or other population differences. The code to implement this model can be found in the supporting information S2.

Of the methods discussed, we would advocate the use of the proportional effects or meta-regression models to incorporate adult data into analyses that estimate the relative effectiveness of treatments in children, as these methods can account for the scenario where children’s responses to medical intervention differs from adults, and this difference can be quantified by parameters estimated from the data. Although not explored in this paper, there are alternative ISMs available that can impose constraints on the relationship between the adult and child population e.g., assuming the relative effectiveness of one population is assumed to be larger or smaller than another or is expected to follow a particular mathematical function, however for these to be considered appropriate in the context of ‘borrowing strength’ from adult data, a substantial amount of previous knowledge and clinical advice would be required to make such assumptions with confidence.

We have shown ISM methods can improve the certainty of clinical effectiveness estimates in the paediatric population. However, we note that that the performance of the methods can differ under different conditions e.g., when used with datasets with different features and different network structures (10). Ultimately, improving the certainty of estimates in children may help to reduce the need for additional RCTs in children, and aid clinicians and patients in making treatment decisions, from the existing evidence base. The models may also be used to facilitate prediction of the effect of new treatments in children (through estimation of a prediction interval) for example, if models such as the ‘proportional effects model’ or ‘meta-regression model’ showed treatment effect estimates of current treatments options were consistent between adults and children, (either consistently similar or had similar differences across comparisons). These predictions could then lead to smaller trials being required to confirm the relative effects of new drugs in children, which could improve the evidence-base in this population and lead to faster approval and uptake of effective treatments.

## Data Availability

All relevant data are within the manuscript and its Supporting Information files.

## Limitations

Here we have focused on simple approaches to information sharing in pairwise meta-analysis, however, other methods have been proposed, namely using the external data as prior information (13) or model averaging approaches (14) to combine the different evidence sources, which we have not evaluated in this paper.

Our work uses an example dataset containing only trials making the same treatment comparison. Collecting data to form a full network of treatments where direct and indirect evidence is available (within populations), could result in stronger evidence and the potential for more information sharing. It would also allow for testing further assumptions e.g., whether relative treatment effects are similar for adults/children across multiple treatments or within a class of treatments but not another.

## Authors roles

RW was responsible for the conceptualization, methodology, data curation, formal analysis, software programming, preparation and presentation of the published work and writing of the original draft for publication.

SD was involved in the conceptualization, methodology, formal data analysis, supervision and reviewing and editing the published work.

BP was involved in the conceptualization, methodology, supervision and reviewing and editing the published work.

## Notes

### Competing Interest Statement

The authors have declared no competing interest.

### Funding Statement

This research was funded by an NIHR Pre-doctoral Fellowship award (Round 1) NIHR300469 held by RW. The views expressed are those of the author(s) and not necessarily those of the NIHR or the Department of Health and Social Care. The funders had no role in study design, data collection and analysis, decision to publish, or preparation of the manuscript.

### Author Declarations

This is a secondary analysis using agreggate data from clincial trial publications and therefore does not require ethical approal.

